# Toward Transportable Acute Kidney Injury Prediction: An Explainable XGBoost Model with Temporal Validation Using MIMIC-IV

**DOI:** 10.64898/2026.09.01.26360393

**Authors:** Daniel Okundaye, Chinyere Chioma Isiekwene

## Abstract

**Background:** Acute kidney injury (AKI) is a frequent complication within intensive care units, with its sudden onset often missed. Existing machine learning and deep learning models have contributed to closing this gap, but their complexity, requiring hundreds to thousands of features, and lack of generalisation pose a limitation that prevents them from being integrated into clinical workflows across different electronic health-record ecosystems.

**Methods:** This study presents a 37-feature XGBoost model trained on the MIMIC-IV dataset with 5.4% positive cases, with hyperparameters optimised via Optuna and probabilities calibrated using isotonic regression, designed for transportability across clinical settings. Validation was conducted internally using a temporal patient-level split simulating prospective deployment, training on 2008–2016 data and testing on 2017–2022 data. External validation was performed on the eICU Collaborative Research Database, a multi-centre dataset spanning 208 US hospitals, using the trained model without retraining. SHAP TreeExplainer was used to provide feature-level explainability for individual predictions.

**Results:** Internal testing yielded an AUROC score of 0.794 for predicting AKI onset within a 12–24 hour window. External validation produced a 0.750 AUROC without retraining. Equitable discrimination was observed across gender, age, chronic kidney disease presence, race, and AKI stages on both datasets, with a 95% internal CI of 0.789–0.799 confirming the model’s estimate stability.

**Conclusion:** These results suggest that clinically useful prediction systems are achievable with substantially fewer features than current models require.

## 1. Introduction

Acute kidney injury (AKI) is the sudden decline in kidney function that affects nearly one in five hospitalised patients across the world, and on its own is associated with increased mortality, prolonged hospital admission windows, and long-term progression to chronic kidney disease [19, 9]. AKI is defined by the Kidney Disease: Improving Global Outcomes (KDIGO) 2012 guidelines as an increase in serum creatinine of 0.3 mg/dL or more within 48 hours, or 1.5 times the baseline value within 7 days, or a sustained reduction in urine output [10]. In intensive care units, the rate of AKI occurrence is higher, with reported cases exceeding 30%, depending on the population and definition applied [5]. Identifying patients at risk early is clinically important because it allows for timely interventions which include nephrotoxic medication review, fluid management, and nephrology referral that can reduce the severity or prevent the onset of kidney injury altogether [13].

Despite the rapid growth of machine learning and its introduction into AKI prediction, the field remains dominated by models built for specific disease populations, and the existing general-population models tend to require many features and integration complexity, limiting their applicability in other ecosystems. Section 2 examines these limitations in detail.

This study presents the development and internal temporal validation of an XGBoost-based AKI prediction model that is designed to address the transportability and interpretability gaps identified in recent systematic reviews. This model makes use of 37 clinical features derived from a general intensive care population obtained from the MIMIC-IV 3.1 database and predicts AKI onset within a 12-to-24-hour window using creatinine-triggered prediction points. An intentional design choice was made to utilise a minimal feature set that relies on routinely collected clinical data, which includes laboratory values, vital signs, demographics, comorbidities, and medication exposure, without needing the hundreds or thousands of features seen in higher-performing models. The model was evaluated using a validation strategy that trained on 2008–2016 data and tested on 2017–2022 data to simulate prospective deployment. Explainability is provided through SHapley Additive Explanations (SHAP) to identify the clinical features driving individual predictions. Code is available upon request.

## 2. Related Work

Machine learning and deep learning have become some of the most promising approaches to timely AKI prediction, with models ranging from logistic regression and gradient boosted trees to deep recurrent neural networks trained on electronic health record (EHR) data. However, the existing literature faces several persistent limitations that restrict the clinical utility of these models on a general scale. Rehman et al. [17] examined 44 externally validated studies composed of 161 AKI prediction models and discovered that many of them were developed in specialised populations, including intensive care, post-surgical, and disease-specific cohorts such as sepsis, heart failure, and liver cirrhosis. Additionally, Wainstein et al. [21] screened 889 studies, specifically searching for externally validated machine learning models built from general hospital populations using the current KDIGO definition and found only three models that met these criteria. An earlier systematic review by Hodgson et al. [4], which evaluated 11 prediction models across general hospital settings that included over 470,000 patient episodes, reported a median AUROC of 0.74 with a high risk of bias in 9 studies, raising concerns around heterogeneous outcome definitions, inadequate calibration reporting, and the problematic use of serum creatinine as both a predictor and an outcome variable.

Even among models with strong results, a recurring problem is that high performance comes at the cost of complexity, reproducibility, and applicability. Tomašev et al. [20] developed a recurrent neural network that was trained on over 703,000 patients across the US Department of Veterans Affairs health system. This model processed around 620,000 features, achieving an AUROC of 0.921 for any-stage AKI prediction within 48 hours. The problem, however, was that the model relied on a predominantly male veteran population with questions raised over its generalisability to other settings due to its dependence on a specific EHR ecosystem. Koyner et al. [12] achieved an AUROC of 0.90 for predicting KDIGO stage 2 or higher AKI only within 24 hours using a gradient boosting machine, and external validation by Churpek et al. [3] maintained strong performance across three health systems. Song et al. [18] demonstrated the issue of cross-site transportability, showing that even high-performing models experienced a degradation when applied to ecosystems with different EHR configurations and demographics, with validation AUROCs ranging from 0.60 to 0.81. Kim et al. [11] developed a recurrent neural network for general inpatients across two Korean tertiary hospitals using 107 features and achieved an AUROC of 0.84 for any-stage AKI and 0.90 for stage 2 or higher, but the model was not tested outside of a single country’s hospital system. Wainstein et al. [21] inferred that this field requires simpler, more transportable prediction models.

Beyond general-population models, disease-specific approaches share similar limitations. Rank et al. [16] developed a recurrent neural network for AKI prediction in post-cardiothoracic surgery patients and achieved an AUROC of 0.893, outperforming expert physicians but remaining limited to that population. Yue et al. [22] applied XGBoost to predict AKI in critically ill patients with sepsis, achieving an AUROC of 0.821, but the model is also restricted to septic patients. Kate et al. [8] built prediction and detection models specifically for hospitalised elderly patients and discovered comorbidities were the strongest predictors of AKI, but these models achieved modest discrimination with AUROCs of 0.66. Mohamadlou et al. [14] developed a gradient boosted tree model using only five bedside measurements and demonstrated prediction 72 hours ahead of onset, but its performance declined at longer prediction horizons. While each of these studies offers useful insights to understanding AKI predictions in different contexts, none of them addressed the need for a general-population model that is simple enough to be transportable, and interpretable for decision-making by clinicians.

With all these put together, the literature reveals a field where strong predictive performance has been demonstrated but at the expense of practical deployment. General-population models remain uncommon, those that exist are complex and difficult to reproduce, disease-specific models do not generalise, and transportability across health systems remains largely unaddressed. This study is positioned as a response to these gaps.

## 3. Methodology

### 3.1 Data Source

This study utilised the Medical Information Mart for Intensive Care IV (MIMIC-IV) version 3.1, a publicly available de-identified clinical database created and maintained by the MIT Laboratory for Computational Physiology and hosted on PhysioNet [7]. MIMIC-IV is composed of electronic health records from Beth Israel Deaconess Medical Center in Boston, Massachusetts, covering admissions and patient-related events from 2008 to 2022. The data includes hospital-level data (diagnoses, laboratory measurements, prescriptions) and ICU-level data (vital signs, fluid inputs and outputs, monitoring records). All dates in the dataset are shifted per patient to preserve privacy, but the temporal ordering across admissions for each patient is maintained. The data was accessed under a signed Data Use Agreement following completion of the Collaborative Institutional Training Initiative (CITI) human subjects research course.

### 3.2 Cohort Definition

The cohort used for the study was obtained from ICU stays within the database. The inclusion criteria required the patients to be 18 years or older and to have a minimum ICU length of stay of 24 hours. Patients below 18 were excluded due to differing paediatric creatinine norm ranges and AKI physiology. Stays shorter than 24 hours were excluded because they lacked a sufficient observation window to label or predict AKI onset. The resulting cohort consisted of 54,551 patients across 74,829 stays.

### 3.3 AKI Labelling

AKI for this model was primarily defined using the serum creatinine criteria of the KDIGO guideline. The criteria used were a creatinine increase of 0.3 mg/dL or more within 48 hours, or a creatinine increase to 1.5 times or more of the baseline value within 7 days. The 48-hour and 7-day baselines were computed as the minimum creatinine values observed within the preceding 48-hour and 7-day windows respectively. Urine output was not used for AKI labelling due to inconsistent recording outside of ICU settings. Creatinine measurements extracted from the *labevents* table were filtered to values greater than 0 and less than 15 mg/dL and restricted to measurements within 24 hours before and after the ICU stay window. AKI staging followed KDIGO: stage 1 for a 7-day ratio of 1.5 or greater, stage 2 for 2.0 or greater, and stage 3 for 3.0 or greater. The cohort exhibited an AKI incidence of 36.1% at the stay level, with 70.6% classified as stage 1, 19.5% as stage 2, and 9.9% as stage 3.

### 3.4 Prediction Point Design

For the study, data was restructured so that each creatinine measurement within an ICU stay served as a prediction point, as opposed to generating a single prediction for the entire stay. This design reflects the reality that the risk of AKI is assessed every time new laboratory results become available. At each prediction point, a binary label of 1 was assigned if AKI onset occurred within 12 to 24 hours after that measurement. Rows where AKI onset was less than 12 hours away were excluded from the dataset, as they fell within a very narrow window for meaningful clinical intervention. All rows at or after the first AKI onset within a stay were deliberately excluded because the model is designed to only predict the occurrence of AKI. Recurrent AKI episodes within the same stay were also not included. This process produced 391,745 prediction points, of which 21,070 (5.4%) carried a positive label.

### 3.5 Feature Engineering

37 features were assigned across seven categories: laboratory values, creatinine trend dynamics, vital signs and trends, temporal context, demographics, comorbidities, and medication exposure.

Creatinine trend dynamics included the current value, 24-hour and 48-hour rolling minimum and maximum, and creatinine change and velocity over both windows. These captured the trajectory of kidney function without exposing the baseline values used in the labelling formula, which were excluded to prevent label leakage.

Other laboratory values were blood urea nitrogen (BUN), sodium, and potassium which were extracted and aligned to each prediction point using a last-observation-carried-forward (LOCF) strategy wherein the most recent value recorded at or before each prediction point was carried forward. The BUN-to-creatinine ratio was computed as a derived feature. Estimated glomerular filtration rate (eGFR) was calculated using the CKD-EPI 2021 race-free equation [6], which was chosen over previous formulations that had race coefficients, to provide equitable kidney function estimates.

Vital signs such as heart rate, peripheral oxygen saturation (SpO_2_), systolic, diastolic, and mean arterial blood pressure, and temperature were collected. Arterial line measurements were preferred for blood pressure where available, with non-invasive cuff measurements used as a fallback to reduce missingness from 64% to approximately 18%. Four additional features were derived: 6-hour rolling minimum and 6-hour change for both mean arterial pressure and SpO_2_.

Temporal features were composed of the number of hours since ICU admission, hours since last creatinine measurement, and the number of creatinine measurements in the previous 24 hours.

Demographics included sex and age; patients aged 89 and older were censored by MIMIC-IV to a single anchor value of 91. This affected 3.3% of the cohort and was noted as a limitation.

Comorbidities were matched with their ICD diagnoses (ICD-9 and ICD-10) and included binary flags for diabetes, hypertension, heart failure, chronic kidney disease, and sepsis.

Medication exposure covered nephrotoxic drugs including NSAIDs, aminoglycosides, vancomycin, and other agents. Medication flags were time-aware and highlighted the presence of a drug at each prediction point based on prescription start and stop times. This produced two features: a binary flag for active nephrotoxic drugs and a count of active nephrotoxic agents.

Missing data was handled according to each feature type. Lab values were carried forward using LOCF, with a binary flag added for potassium measurement due to 58% missing proportion. Comorbidity flags were zero-filled, indicating absence of a condition. The remaining missing values in laboratory and vital sign features were left as NaN, as XGBoost naturally handles missing values.

### 3.6 Model Training

The primary model was XGBoost, a gradient boosted tree algorithm that was selected for its established performance on tabular clinical data and its ability to handle missing values without imputation. The class imbalance was addressed using the `scale_pos_weight` parameter, set to 17.526 (ratio of negative to positive training examples), which increases the loss function of missed positive cases without generating synthetic data.

Hyperparameter optimisation was done via Optuna tuning with 50 trials and a Tree-structured Parzen Estimator (TPE) sampler. Each trial was evaluated using 3-fold GroupKFold cross-validation on the training set, with groups defined at the patient level to ensure that all prediction points from a given patient appeared in the same fold. This prevented the model from seeing a patient’s clinical patterns during training and validation in the same cross-validation loop. The optimal hyperparameters were `max_depth` = 6, `learning_rate` = 0.013, `subsample` = 0.766, `n_estimators` = 851, `colsample_bytree` = 0.656, `reg_alpha` = 2.477, `reg_lambda` = 5.573, and `min_child_weight` = 4.

A logistic regression baseline model was trained using median imputation and standard scaling to provide a performance floor for comparison.

### 3.7 Temporal Validation

The model was evaluated using a temporal patient-level split that aligned with the `anchor_year_group` boundaries in MIMIC-IV to simulate prospective deployment. Training data consisted of ICU stays from 2008–2016 (272,705 prediction points, 14,720 positive cases) and test data consisted of ICU stays from 2017–2022 (119,040 prediction points, 6,350 positive cases). Patient-level split ensures that all stays from a given patient appeared either in the training set or test set.

### 3.8 Calibration

Post-hoc calibration was applied via isotonic regression with `CalibratedClassifierCV` with 3-fold cross-validation. This was necessary due to the `scale_pos_weight` parameter inflating predicted probabilities, even though discrimination was improved. Isotonic regression was chosen over Platt scaling because the miscalibration pattern was not S-shaped. Calibration yielded probabilities that were compressed into a range that closely reflected the true 5.4% positive rate, while AUROC was preserved.

### 3.9 Evaluation

Model performance was assessed using AUROC, area under the precision-recall curve (AUPRC), and the Brier score. Three operating points were reported: a high-specificity threshold, the Youden threshold (which maximises sensitivity plus specificity minus one), and a high-sensitivity threshold. A confusion matrix and F1 score were computed at the Youden threshold and SHAP TreeExplainer was applied to a 1,000-row random sample of the test set to identify the most influential features in the model’s predictions. Subgroup analysis was conducted across sex, age, race, and CKD status to assess model equity. Lastly, bootstrap confidence intervals (95%) were computed using 1,000 iterations with replacement on the test set.

### 3.10 External Validation

External validation was conducted on the eICU Collaborative Research Database [15], a multi-centre dataset comprising over 200,000 ICU admissions from 208 hospitals across the United States during 2014–2015. The validation cohort was constructed using the same inclusion criteria for internal training and testing on the MIMIC-IV cohort: adults aged 18 or older with ICU stays of at least 24 hours, yielding 110,257 stays across 95,823 patients. The same feature pipeline was rebuilt using eICU’s schema, with lab values identified by name rather than ID and temporal offsets in minutes rather than datetime. 588,274 prediction points were produced with a 3.6% positive rate. Missing vital signs were considerably higher in eICU, with blood pressure and temperature missing approximately 77% and 89% of prediction points respectively, compared to 18% and 20% in MIMIC-IV. The calibrated XGBoost model was applied directly to the feature matrix without retraining or additional calibration.

## 4. Results

### 4.1 Model Progression

Four model configurations were assessed to establish a performance trajectory and discover the source of predictive signal. Table 1 summarises the progression from a logistic regression baseline model to the final tuned and calibrated XGBoost model. LightGBM was evaluated as an alternative algorithm on the same feature set and achieved an AUROC of 0.793, confirming that the performance ceiling was driven by the available features rather than the choice of algorithm. This conclusion was further supported with the separate inclusion and evaluation of urine output features which contributed less than 0.001 AUROC.

**Table 1.** Model progression across development stages.

| Model | Feat. | AUROC | AUPRC |
| --- | --- | --- | --- |
| Logistic Regression | 27 | 0.755 | 0.167 |
| XGBoost (default) | 27 | 0.771 | 0.191 |
| XGBoost (tuned, cal.) | 27 | 0.789 | 0.208 |
| **XGBoost (tuned, cal.)** | **37** | **0.794** | **0.213** |
| LightGBM (tuned) | 33 | 0.793 | 0.210 |

### 4.2 Final Model Performance

The final calibrated XGBoost model achieved the performance metrics shown in Table 2. The Brier score of 0.046 compared favourably against a baseline of 0.051 for a model predicting the mean positive rate for all examples.

**Table 2.** Final model metrics with bootstrap 95% CIs (1,000 iterations).

| Metric | Value | 95% CI |
| --- | --- | --- |
| AUROC | 0.794 | 0.789–0.799 |
| AUPRC | 0.213 | 0.203–0.222 |
| Brier Score | 0.046 | — |

Three operating points were identified and are presented in Table 3. At the Youden threshold (0.044), the model achieved an F1 score of 0.218 with a precision of 12.8% and recall of 74.8%. The confusion matrix at this threshold is shown in Table 4. The low precision is a direct consequence of the 5.4% positive rate; even with good discrimination, the large number of negative cases produces many false positives at any threshold that achieves reasonable sensitivity, resulting in approximately 7 false alarms for every true AKI alert.

**Table 3.** Operating points on the temporal test set.

| Point | Thresh. | Sens. | Spec. |
| --- | --- | --- | --- |
| High Specificity | 0.067 | 62.1% | 80.0% |
| Youden | 0.044 | 74.8% | 70.0% |
| High Sensitivity | 0.036 | 80.0% | 63.2% |

**Table 4.** Confusion matrix at Youden threshold (0.044).

|  | Pred. Neg. | Pred. Pos. |
| --- | --- | --- |
| Actual Negative | 81,502 (TN) | 31,188 (FP) |
| Actual Positive | 1,757 (FN) | 4,593 (TP) |

**Table 5.** Decision curve analysis summary.

| Threshold Range | Model vs Treat All | Model vs Treat None |
| --- | --- | --- |
| 0.01–0.05 | Marginal advantage | Model superior |
| 0.05–0.20 | Model clearly superior | Model superior |
| 0.20–0.35 | Model superior | Marginal advantage |
| > 0.35 | Both near zero | Both near zero |

The model underwent decision curve analysis to evaluate whether using the model to guide clinical decisions provides net benefit over default strategies (treat all and treat none). Figure 4 illustrates that the model performed better than the default strategies across thresholds of 0.01 to 0.35, with stronger performance within the 0.05–0.20 range, which aligns with the three operating points determined. “Treat all”, which represents the event where all patients are treated, dropped below zero at a threshold of 0.08, highlighting that treating everyone causes more harm than benefit from unnecessary interventions.

**Figure 1.**
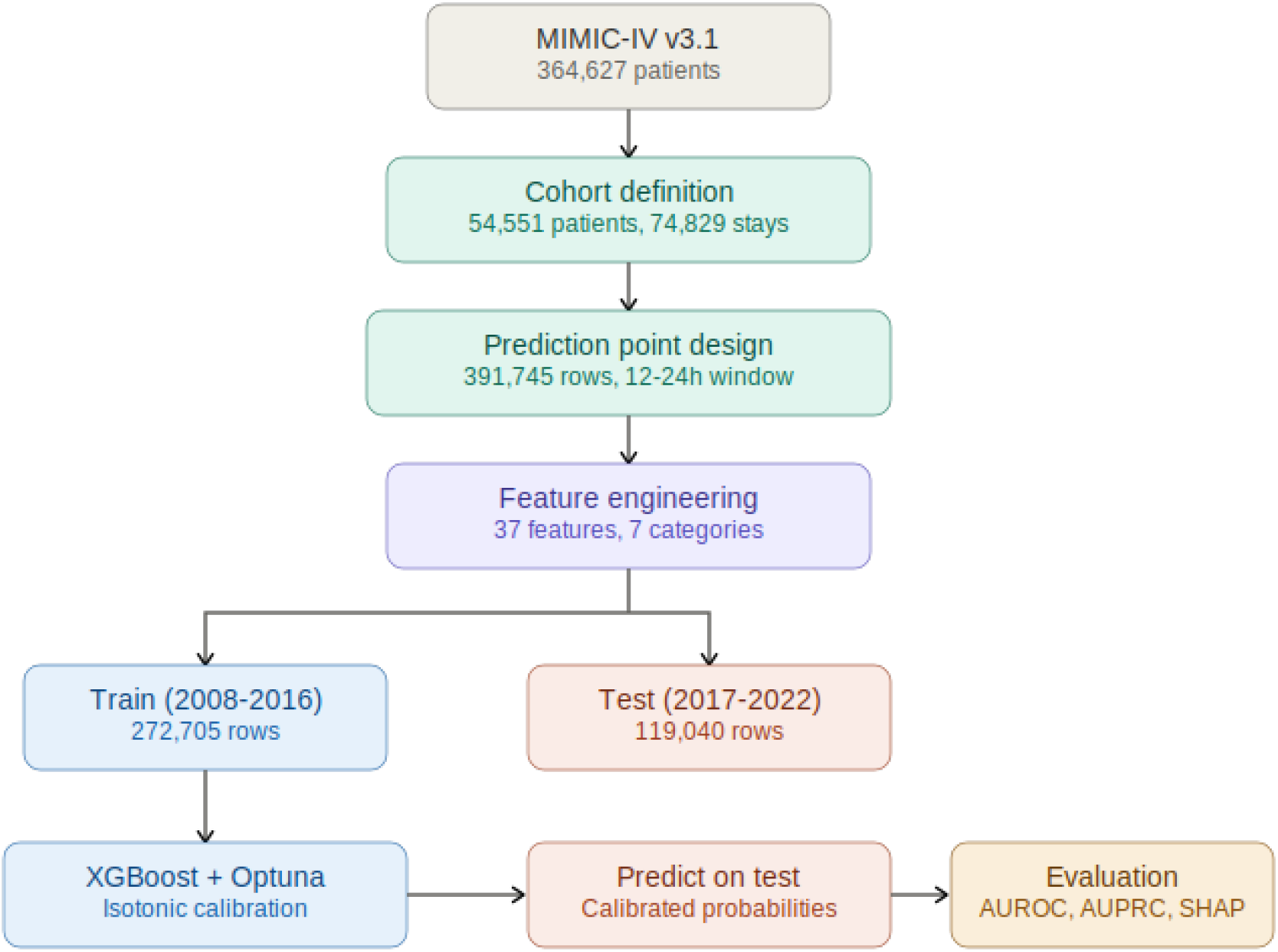
Pipeline flowchart illustrating the data processing, feature engineering, model training, and evaluation workflow.

**Figure 2.**
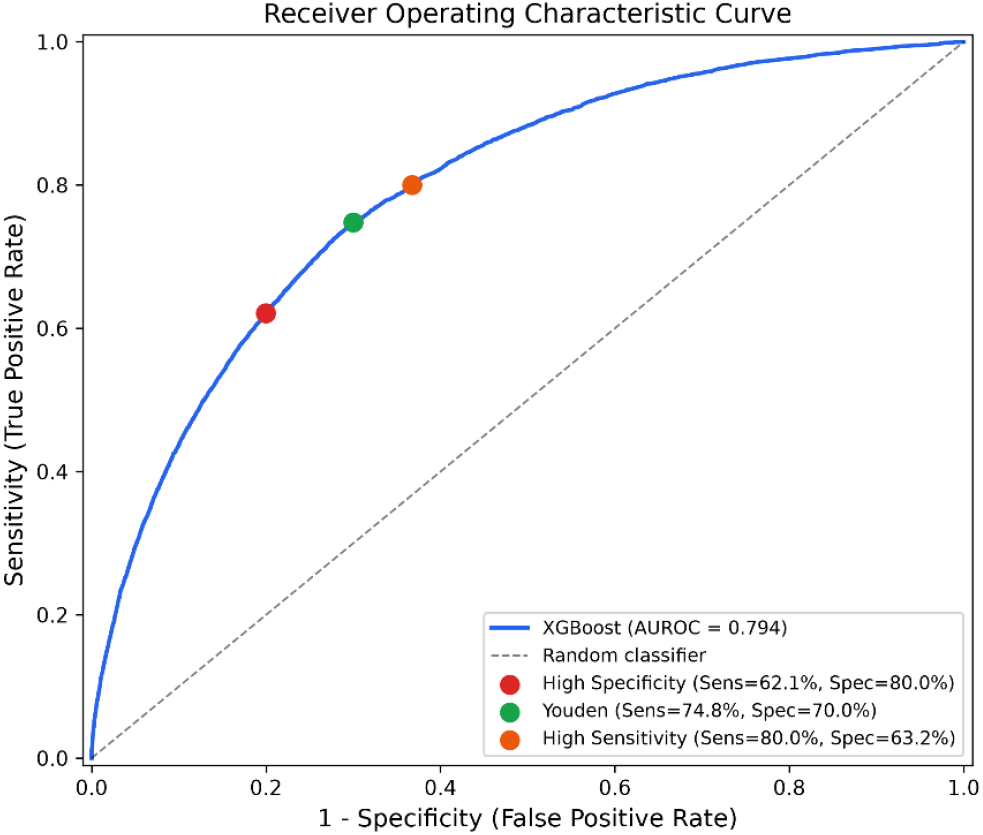
Receiver operating characteristic (ROC) curve on the temporal test set.

**Figure 3.**
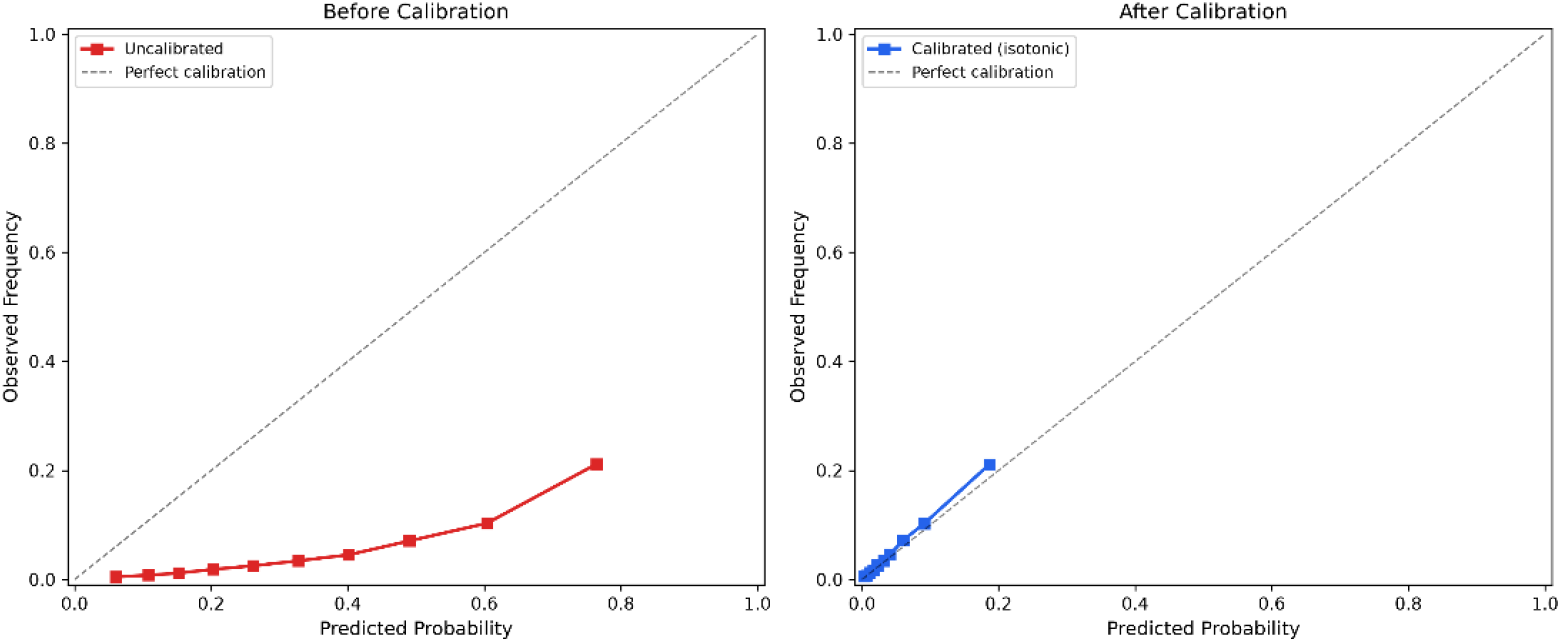
Calibration curve comparing predicted probabilities against observed AKI rates.

**Figure 4.**
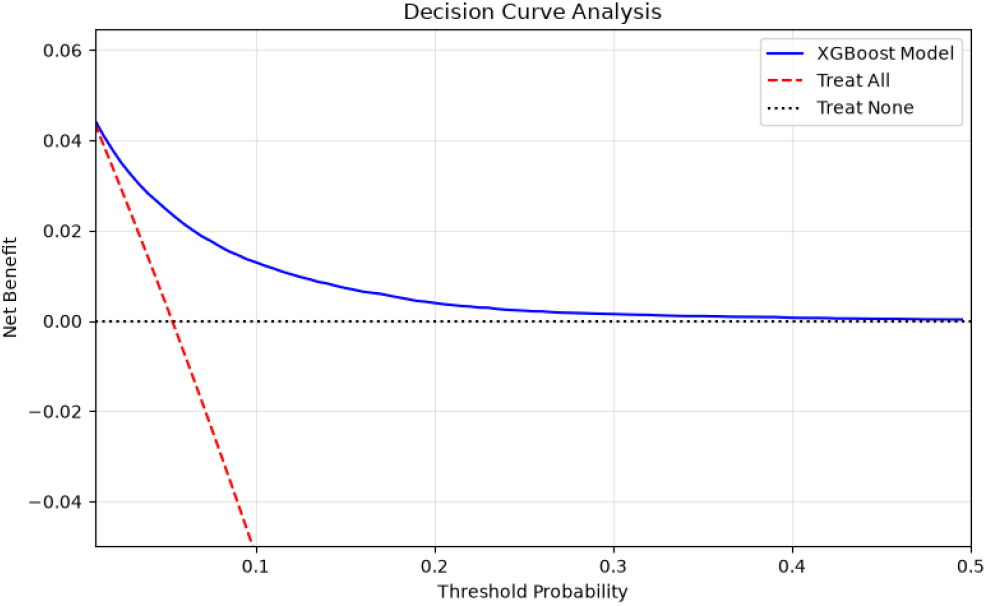
Decision curve analysis comparing net benefit of the model against default strategies.

**Figure 5.**
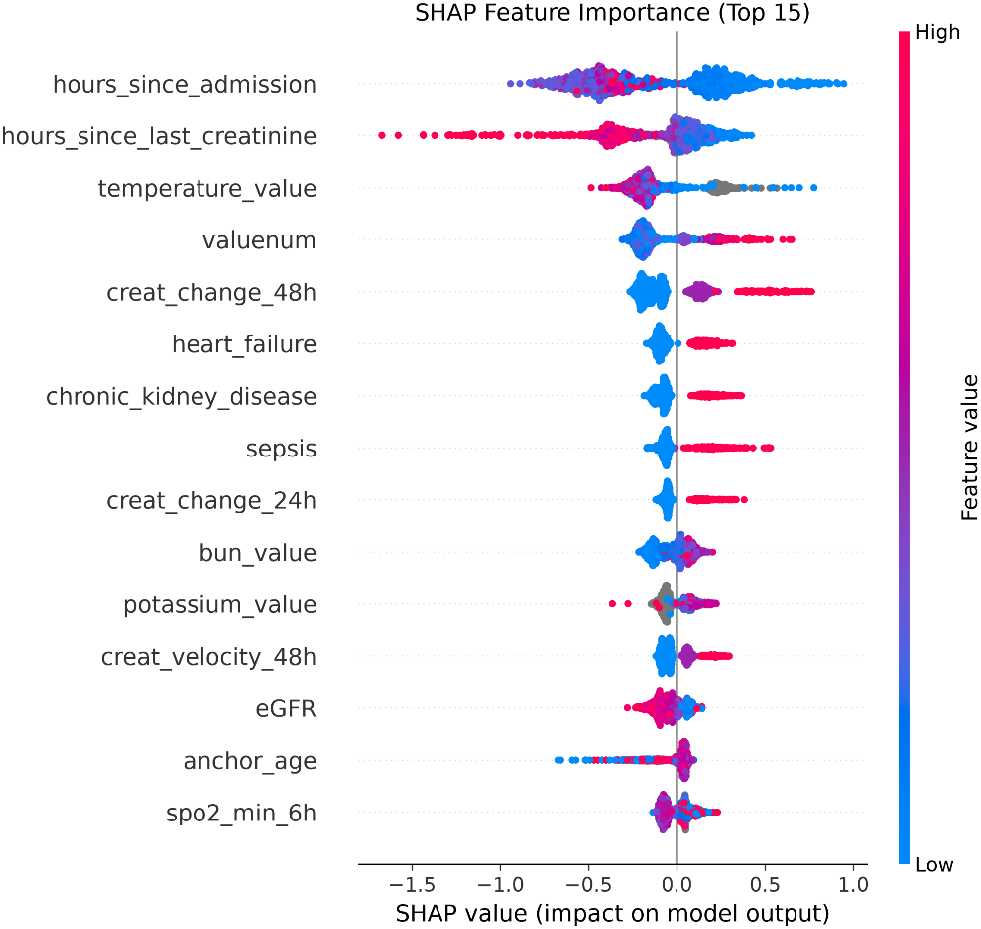
SHAP summary plot showing feature importance and directional impact on predictions.

### 4.3 Feature Importance

Application of the SHAP TreeExplainer analysis on a 1,000-row test sample identified the features shown in Table 6 as the most influential. Temporal features dominated, with hours since ICU admission ranking first. Temperature ranked unexpectedly high, with low temperatures (hypothermia) associated with increased AKI risk, consistent with severe sepsis presentations. Nephrotoxic medication features ranked near the bottom of the importance list.

**Table 6.** Top 10 SHAP features by mean absolute SHAP value.

| Rank | Feature | Category |
| --- | --- | --- |
| 1 | Hours since admission | Temporal |
| 2 | Hours since last creatinine | Temporal |
| 3 | Temperature | Vital |
| 4 | Current creatinine | Lab |
| 5 | Creatinine change (48h) | Creat. trend |
| 6 | Heart failure | Comorbidity |
| 7 | Chronic kidney disease | Comorbidity |
| 8 | Sepsis | Comorbidity |
| 9 | Creatinine change (24h) | Creat. trend |
| 10 | BUN | Lab |

### 4.4 Subgroup Analysis

Model performance was assessed across clinically relevant subgroups to evaluate equity and assess potential bias. Results are presented in Tables 7 and 8.

**Table 7.** AUROC by demographic and clinical subgroup.

| Subgroup | AUROC | Delta |
| --- | --- | --- |
| Female | 0.803 | — |
| Male | 0.786 | 0.017 |
| Age < 65 | 0.793 | — |
| Age ≥ 65 | 0.792 | 0.001 |
| No CKD | 0.779 | — |
| CKD | 0.762 | 0.017 |

**Table 8.** AUROC by AKI severity and race.

| Subgroup | AUROC | N (test) | Positive Cases |
| --- | --- | --- | --- |
| Stage 1 vs No AKI | 0.799 | — | 4,359 |
| Stage 2+ vs No AKI | 0.783 | — | 1,991 |
| White | 0.798 | 65,432 | 3,443 |
| Black/African American | 0.781 | 6,295 | 414 |
| Asian | 0.794 | 1,436 | 58 |
| Hispanic/Latino (Puerto Rican) | 0.812 | 1,414 | 90 |
| Asian (Chinese) | 0.763 | 1,261 | 72 |
| Hispanic/Latino (Dominican) | 0.720 | 1,227 | 61 |
| Unknown/Unable to Obtain | 0.787 | 26,175 | 1,384 |

The largest, reliable gap across race was between White (0.798) and Black or African American (0.781) patients at 0.017. The Hispanic/Latino Dominican subgroup showed the lowest AUROC (0.720) but had only 61 positive cases, making the estimate unreliable.

### 4.5 External Validation

External validation was carried out on the eICU dataset without model retraining. Table 9 presents the discrimination and calibration metrics as the AUROC decreased from 0.794 on MIMIC-IV to 0.750 on eICU with a drop of 0.044. Subgroup analysis on eICU is shown in Table 10, as no subgroup fell below an AUROC of 0.73. The gender gap reversed direction between datasets, and this suggests that gender-related differences are population dependent rather than model bias.

**Table 9.** External validation metrics on eICU (208 US hospitals, no retraining).

| Metric | MIMIC-IV | eICU | Delta |
| --- | --- | --- | --- |
| AUROC | 0.794 | 0.750 | 0.044 |
| AUPRC | 0.213 | 0.125 | 0.088 |
| Brier Score | 0.046 | 0.034 | — |
| Prediction Points | 119,040 | 588,274 | — |
| Positive Cases | 6,350 (5.4%) | 20,986 (3.6%) | — |
| Hospitals | 1 | 208 | — |

**Table 10.** AUROC by subgroup on eICU external validation.

| Subgroup | eICU AUROC | MIMIC-IV AUROC | N | Positive Cases |
| --- | --- | --- | --- | --- |
| Female | 0.741 | 0.803 | 271,227 | 9,384 |
| Male | 0.757 | 0.786 | 317,047 | 11,602 |
| Age < 65 | 0.752 | 0.793 | 284,046 | 9,454 |
| Age ≥ 65 | 0.746 | 0.792 | 304,228 | 11,532 |
| No CKD | 0.735 | 0.779 | 543,288 | 17,301 |
| CKD | 0.754 | 0.762 | 44,986 | 3,685 |
| Caucasian | 0.744 | 0.798* | 452,956 | 15,222 |
| African American | 0.779 | 0.781* | 65,779 | 2,932 |
| Hispanic | 0.734 | — | 21,423 | 909 |
| Asian | 0.740 | 0.794* | 10,602 | 402 |
| Native American | 0.755 | — | 3,426 | 147 |
| Other/Unknown | 0.762 | 0.787* | 28,340 | 1,170 |

\*MIMIC-IV race categories differ from eICU. Comparisons are approximate.

## 5. Discussion

### 5.1 Performance in Context

The final configured model achieved an AUROC of 0.794 on temporal validation, which is lower than several published AKI prediction models including Tomašev et al. [20] (0.921), Koyner et al. [12] (0.90), and Churpek et al. [3] (0.85–0.86). However, direct comparison (Table 11) requires acknowledging the fundamental differences in model design. Tomašev etal. [20] processed approximately 620,000 features through a deep recurrent neural network, Koyner et al. [12] predicted only stage 2 or higher AKI, and Churpek et al. [3] used 59 features with continuous prediction across emergency department, ward, and ICU settings. A recent analysis conducted by Cama-Olivares et al. [1] pooled 150 studies with a total of 14.4 million participants reported an external validation AUROC of 0.78, with 86% of the studies carrying a high risk of bias per PROBAST. This model employs 37 features and a simpler gradient boosted tree architecture, deliberately trading peak discrimination for a design that prioritises transportability and interpretability. It also maintains methodological safeguards including calibration, bootstrap confidence intervals, and subgroup equity analysis that most published models omit.

**Table 11.**
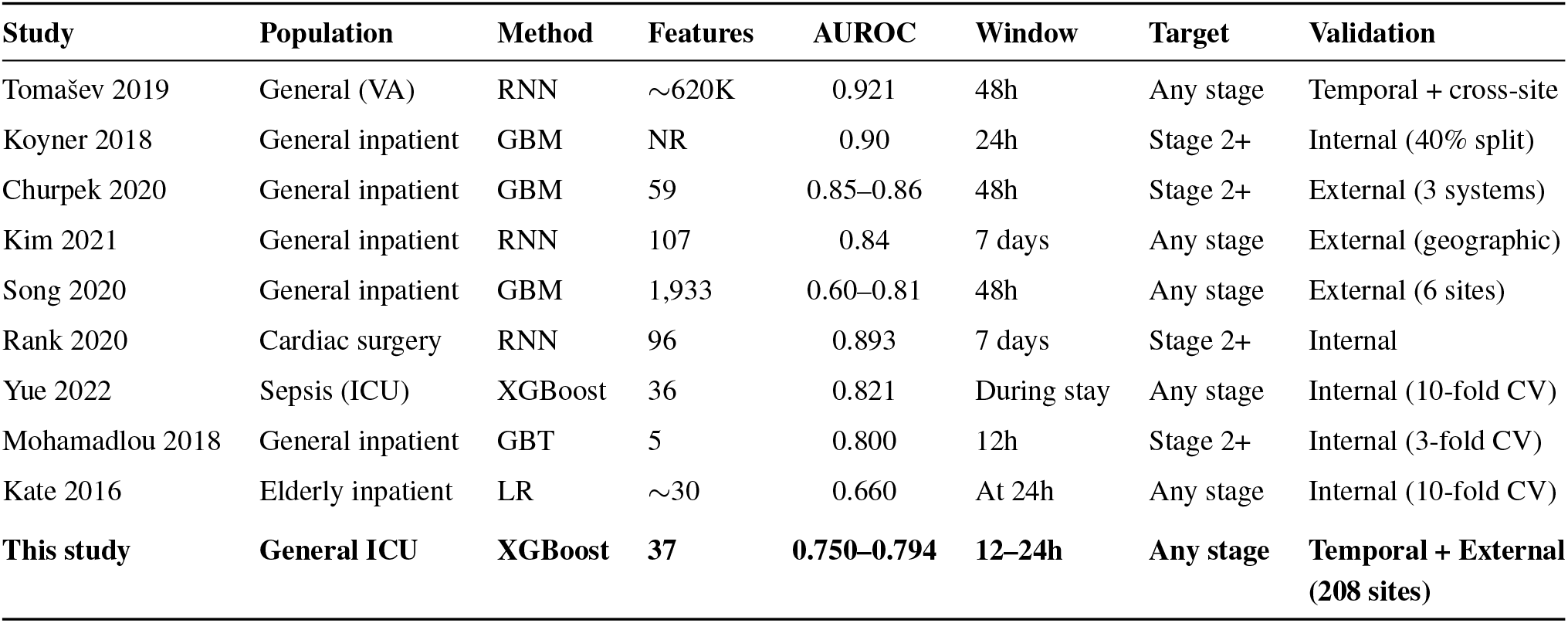
Comparison with published AKI prediction models.

The model’s AUROC is consistent with the range reported by Song et al. [18] (0.60–0.81) in their cross-site transportability study, where performance varied substantially depending on the target site’s EHR configuration and patient demographics. It also exceeds the median AUROC of 0.74 reported across 11 general-population models in the systematic review by Hodgson et al. [4] and falls within the range of the three externally validated models identified by Wainstein et al. [21].

External validation on eICU (208 hospitals) yielded an AUROC of 0.750 without retraining, showcasing a 0.044 decrease from internal validation. This degradation is substantially smaller than the range reported by Song et al. [18] and supports the claim that a minimal-feature design reduces cross-site performance loss. Higher vital sign absence in eICU in comparison to MIMIC-IV likely contributed to the performance drop.

### 5.2 Feature-Driven Performance Ceiling

A notable finding of this study is that the performance ceiling was driven by the available features rather than the algorithm used as seen in both XGBoost and LightGBM achieving near-identical AUROCs (0.794 and 0.793 respectively) on the same feature set, and adding vital sign trends produced minimal improvement. Urine output features contributed less than 0.001 AUROC, suggesting that creatinine trajectory, temporal context, and comorbidity features already capture the predictive signal available in the data, and that further improvement requires very different feature sources such as clinical notes or richer medication data including dosing and administration.

This finding aligns with the observation by Wainstein et al. [21] that serum creatinine and its derivatives were consistently among the top predictors in all three models reviewed, raising the question of what these models offer beyond what creatinine measurements alone provide. In this model, temporal features (hours since admission and hours since last creatinine) ranked above creatinine itself, suggesting that monitoring intensity and care patterns carry independent predictive signal. Temperature’s high ranking (third overall) also provides a non-creatinine signal, with hypothermia likely reflecting severe sepsis or haemodynamic instability.

### 5.3 Subgroup Equity

The model demonstrated minimal performance variation across key demographic subgroups. The largest delta was 0.017 between female and male patients, and between White and Black or African American patients. The age delta was 0.001 and CKD delta was also 0.017, highlighting that these gaps are small relative to those reported in other clinical prediction models and suggest reasonable equity across the sub-groups assessed. The use of the CKD-EPI 2021 race-free equation for eGFR calculation [6] was a deliberate design choice to avoid embedding racial bias into kidney function estimation.

The model predicted stage 1 AKI slightly better than stage 2 or higher (0.799 versus 0.783). This either reflects the dominance of stage 1 in the training data (70.6% of all AKI cases) or the tendency of severe AKI to develop rapidly from acute events such as haemorrhage or septic shock, where the creatinine trajectory before onset does not follow the progressive pattern the model learned from more common mild cases.

The low precision at the Youden threshold (12.8%, or approximately 1 true alert for every 7 false alarms) is a direct consequence of the 5.4% positive rate and is consistent with the alert fatigue challenge documented across AKI prediction systems. Operating point selection in deployment would need to balance sensitivity against the clinical capacity to respond to alerts.

### 5.4 Transportability and Reproducibility

Wainstein et al. [21] identified three critical barriers to the practical deployment of AKI prediction models: excessive feature requirements, dependence on specific EHR ecosystems, and lack of code availability. This study was designed to address each of these concerns by using 37 features drawn from routinely collected clinical data without requiring specialised variables or hospital-specific data fields. External validation on the eICU Collaborative Research Database which spans 208 hospitals across the United States with different EHR integrations yielded 0.750 AUROC. Its performance drop of 0.044 is consistent with the expected cross-site degradation reported by Song et al. [18] but remains substantially smaller than the range of 0.60–0.81. Code for the full pipeline from data processing through model training and evaluation is available upon request to support reproducibility.

### 5.5 Future Work

A two-model architecture was originally conceived for this system: the lab-based predictor presented here (Model B) and a symptom-based outpatient screener (Model A) designed for patients without laboratory results. Model A would use demographics, presenting symptoms, medical history, and medication exposure to identify patients who should be referred for kidney function testing. This would have addressed a gap identified across the literature, where no existing model targets outpatient populations without lab data. Cheng et al. [2] developed the closest comparator, but it still required laboratory values as inputs. Development of Model A requires prospective symptom data collection from a clinical setting and remains a planned extension of this work.

Integration with electronic medical record systems via the FHIR R4 standard is also planned. A FHIR-compliant data pipeline would allow the model to receive patient data as standardised resources, reducing the EHR-specific data formatting that Song et al. [18] identified as a source of cross-site performance degradation.

### 5.6 Limitations

This study has several limitations that should be considered when interpreting the results.

First, although external validation demonstrated reasonable cross-site performance, both MIMIC-IV and eICU are US-based datasets. The model has not been validated on non-US healthcare systems or community hospitals with limited monitoring capabilities or outpatient settings that differ from trained ICU environments.

Second, the model was trained on ICU patients who have dense monitoring and frequent laboratory measurements. Deploying this model in ward or outpatient settings, where measurement frequency is lower and missingness patterns differ, may result in performance degradation. The LOCF imputation strategy assumes a recent prior value exists, which may not hold outside the ICU.

Third, AKI labelling relied exclusively on serum creatinine criteria. The urine output criterion of the KDIGO definition was excluded due to inconsistent recording. This may undercount AKI cases, particularly those presenting initially with oliguria before creatinine elevation.

Fourth, MIMIC-IV censors patients aged 89 and older to a single anchor age value of 91, affecting 3.3% of the cohort. The model treats all patients in this group as having identical age, which may underestimate risk for the oldest patients.

Fifth, the creatinine-triggered prediction design means the model only generates predictions when a new creatinine measurement is available. Patients who are not having creatinine monitored would not receive predictions, introducing a selection bias toward patients already under clinical suspicion.

## 6. Conclusion

Approaches to acute kidney injury prediction face a persistent trade-off between performance and accessibility. The more complex a model is, the more strictly it is tied to a specific population or healthcare ecosystem. This model helps to bridge these gaps by building a minimal-feature system without specialised variables allowing it to potentially be deployed across different healthcare systems. This is demonstrated through its achievement of an AUROC of 0.794 on internal validation and 0.750 on external validation across 208 US hospitals, without retraining. The narrow confidence interval (95% CI: 0.789–0.799) and external validation suggest that clinically useful AKI prediction is achievable without the hundreds or thousands of features that characterise higher-performing but less transportable models. Future improvements will pursue validation on non-US datasets and explore the integration of additional features such as clinical notes through NLP, urine output criteria, and deployment evaluation in a real-world setting.

## Data Availability

This study utilized two publicly available de-identified datasets accessed through PhysioNet with signed Data Use Agreements: MIMIC-IV v3.1 and eICU Collaborative Research Database v2.0. Both require completion of CITI human subjects training and PhysioNet credentialing. Code is available upon request from the corresponding author.

https://physionet.org/content/mimiciv/3.1/

https://physionet.org/content/eicu-crd/2.0/

## Notes

### Competing Interest Statement

The authors have declared no competing interest.

### Author Declarations

The Ethics Committee of the School of Computing, Miva Open University gave ethical approval for this work.

